# Fully implanted adaptive deep brain stimulation in freely moving essential tremor patients

**DOI:** 10.1101/2020.05.28.20092148

**Authors:** BI Ferleger, B Houston, MC Thompson, SS Cooper, KS Sonnet, AL Ko, JA Herron, HJ Chizeck

## Abstract

**Objective:** Deep brain stimulation (DBS) is a safe and established treatment for essential tremor (ET) and several other movement disorders. One approach to improving DBS therapy is adaptive DBS (aDBS), in which stimulation parameters are modulated in real time based on biofeedback from either external or implanted sensors. Previously tested systems have fallen short of translational applicability due to the requirement for patients to continuously wear the necessary sensors or processing devices, as well as privacy and security concerns.

**Approach:** We designed and implemented a translation-ready training data collection system for fully implanted aDBS. Two patients chronically implanted with electrocorticography strips over the hand portion of M1 and DBS probes in the ipsilateral ventral intermediate nucleus of the thalamus for treatment of ET were recruited for this study. Training was conducted using a translation-ready distributed training procedure, allowing a substantially higher degree of control over data collection than previous works. A linear classifier was trained using this system, biased towards activating stimulation in accordance with clinical considerations.

**Main Results:** The clinically relevant average false negative rate, defined as fraction of time during which stimulation dropped below 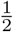 clinical levels during movement epochs, was 0.036. Tremor suppression, calculated through analysis of gyroscope data, was 33.2% more effective on average with aDBS than with continuous DBS. During a period of free movement with aDBS, one patient reported a slight paresthesia; patients noticed no difference in treatment efficacy between systems.

**Significance:** Here is presented the first translation-ready training procedure for a fully embedded aDBS control system for MDs and one of the first examples of such a system in ET, adding to the consensus that fully implanted aDBS systems are sufficiently mature for broader deployment in treatment of movement disorders.

## 1. Introduction

Essential tremor (ET) is the world’s most common type of movement disorder (MD), affecting an estimated 4.6% of the population over age 65 [1]. ET is characterized by its cardinal symptoms of kinetic and postural upper limb tremor, as opposed to the rest tremor common in other conditions [2, 3]. Deep brain stimulation (DBS) of the ventral intermediate nucleus (VIM) of the thalamus is an established treatment for ET [1, 4, 5, 6]. Follow up studies have demonstrated that DBS is a safe and effective treatment for ET, with most adverse effects related to perioperative complications and occurring at low rates [7, 8, 9]. At present, continuous, or conventional, DBS (cDBS), wherein stimulation parameters are set by a clinician and left continuously at those levels, is the standard of clinical care and has been chronically implanted in an estimated 160,000 patients as of 2019 [10].

Nonetheless, cDBS remains an imperfect treatment. The implantable pulse generator (IPG) battery may only be replaced through revision surgeries that must be conducted every few years. Several side effects, including paresthesia, difficulty speaking, balance issues, and sexual emotional disinhibition, are associated with cDBS treatment [11, 12, 13].

One approach to ameliorating these concerns is adaptive DBS (aDBS), interchangeably referred to as closed-loop DBS. Using external or implanted sensors to provide biofeedback, aDBS uses inferences about the patient’s state extracted from this sensed data to modulate stimulation parameters and reduce overall stimulation [14, 15, 16, 17, 18]. This has been shown to dramatically reduce battery drain while potentially reducing the incidence and severity of side effects [14, 17, 18, 19, 20]. The unique nature of ET among conditions for which DBS is prescribed – specifically, the predictable and activity-dependent nature of symptom manifestation – makes it especially well-suited as a testbed for the investigation of aDBS strategies.

Most previously investigated aDBS systems have made use of either external sensing methods or distributed data processing structures. External sensing, such as inertial measurement unit (IMU) data from a smartwatch affixed to the patient’s treated wrist, provides easily defined and understood real-time feedback on symptom severity and patient activity [17]. Distributed data processing structures use data from implanted sensors, such as local field potential (LFP) data recorded using electrocorticography (ECoG) strips, in order to infer the patient’s state through the deployment of machine learning algorithms on an external experimental computer or other computational device [16, 20, 21, 22]. The use of a distributed system allows the application of powerful data processing and machine learning techniques on notoriously noisy, non-stationary neural data. This permits development of highly accurate algorithms to predict patient state.

However, each of these system architectures entails significant drawbacks. External sensing systems require a patient to wear some symptom-tracking device at virtually all times, which is unlikely to serve in a translational capacity due to the likelihood of patients either forgetting the required device some days, or even their unwillingness to wear it altogether. Likewise, distributed systems tether patients to an associated data processing device, limiting mobility in cases where the tether is a physical wire and, with wireless systems, running into the same translational problems encountered with wearable devices [23]. Additionally, both wearable and distributed systems raise clear privacy and security concerns due to the streaming of protected personal information and, perhaps more concerningly, the increased potential for malicious third-party interference with the device itself introduced by this streaming [24].

A clear solution to these concerns is a fully implanted system, in which internally detected biomarkers are processed in the IPG itself and an on-board algorithm applied to modulate stimulation parameters in real time[18]. This system would eliminate concerns about a patient remembering their external devices by making the process entirely internal and automated, with the added benefit of intrinsically resolving issues of privacy and security during chronic treatment. However, due to the necessity of using only internally recorded data and the limited processing power available on implanted devices relative to modern computers and mobile devices, these systems would lack the ease of interpretability in programming ensured by wearable systems and the sheer processing power available to distributed systems. This, in turn, has previously limited the translational applicability of fully implanted aDBS.

Here is presented the first translation-ready training procedure for a fully embedded aDBS control system for use in MDs, as well as one of the first examples of fully embedded aDBS in ET. Through a straightforward distributed training procedure permitting reliable collection of training data, a supervised machine learning algorithm optimized for clinical considerations was used to train intrinsically personalized aDBS classifiers in two patients. These classifiers were then uploaded to the patients’ devices and their performance evaluated through IMU data analysis and patient feedback. This system greatly simplifies the training procedure itself while simultaneously increasing reliability and transparency of classifier training.

## 2. Methods and Materials

### 2.1. Subject information and device specifications

Two subjects diagnosed with ET were implanted with the Activa PC+S, an FDA approved investigational neurostimulator developed by Medtronic with a DBS lead implanted unilaterally in the VIM thalamus and an ECoG-sensing strip of electrodes placed over the hand portion of the ipsilateral motor cortex. These subjects, hereafter referred to as Patient 1 (P1) and Patient 2 (P2), were enrolled in ongoing studies at the University of Washington Medical Center under IRB supervision. Patient 1 is an 84-year-old right-handed male and Patient 2 is a 70-year-old right-handed male.

The on-board classifier of the Activa PC+S is limited to using a bandpower estimate of a given band of LFP data, calculated in a proprietary hardware design. The band must be of the form *c* ± *r* with *c* = {2.5, 5, 7.5 … 97.5, 100}Hz and *r* = {2.5, 8, 16}Hz, with an estimate generated every 200ms (5Hz). Two different bandwidths may be collected from the DBS probe and the ECoG strip, respectively, resulting in a state estimate 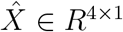 defined as

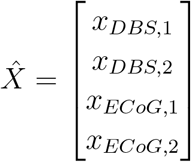

Furthermore, due to the limited processing capacity of the IPG itself, we are restricted to a binary linear classifier to determine whether stimulation amplitude should be set to a pre-determined “LOW” value (set in this work to 0.0V in both patients) or “HIGH” value (a clinician-determined, patient-specific amplitude), with other parameters (frequency, pulse width, stimulation electrode configuration) held constant.

### 2.2. Distributed training architecture

Despite the streaming capacity of the Activa PC+S, previous efforts towards using the device’s fully embedded aDBS capacity required the researcher to rely solely on the on-board capacity of the device to record data in order to obtain training data [18]. This data could then be downloaded to a proprietary tablet, from which it had to be transported by USB to an experimental computer for analysis. In contrast, our system is structured so that training data is directly streamed from the IPG to the experimental computer. This allows for accurate and reliable timestamped data collection, and permits the instantaneous review of all training data to easily determine if a test must be repeated. When satisfactory results have been obtained, the classifier is uploaded to the IPG itself for evaluation of aDBS in free movement. A diagram of this system may be found in fig. 1.

**Figure 1.**
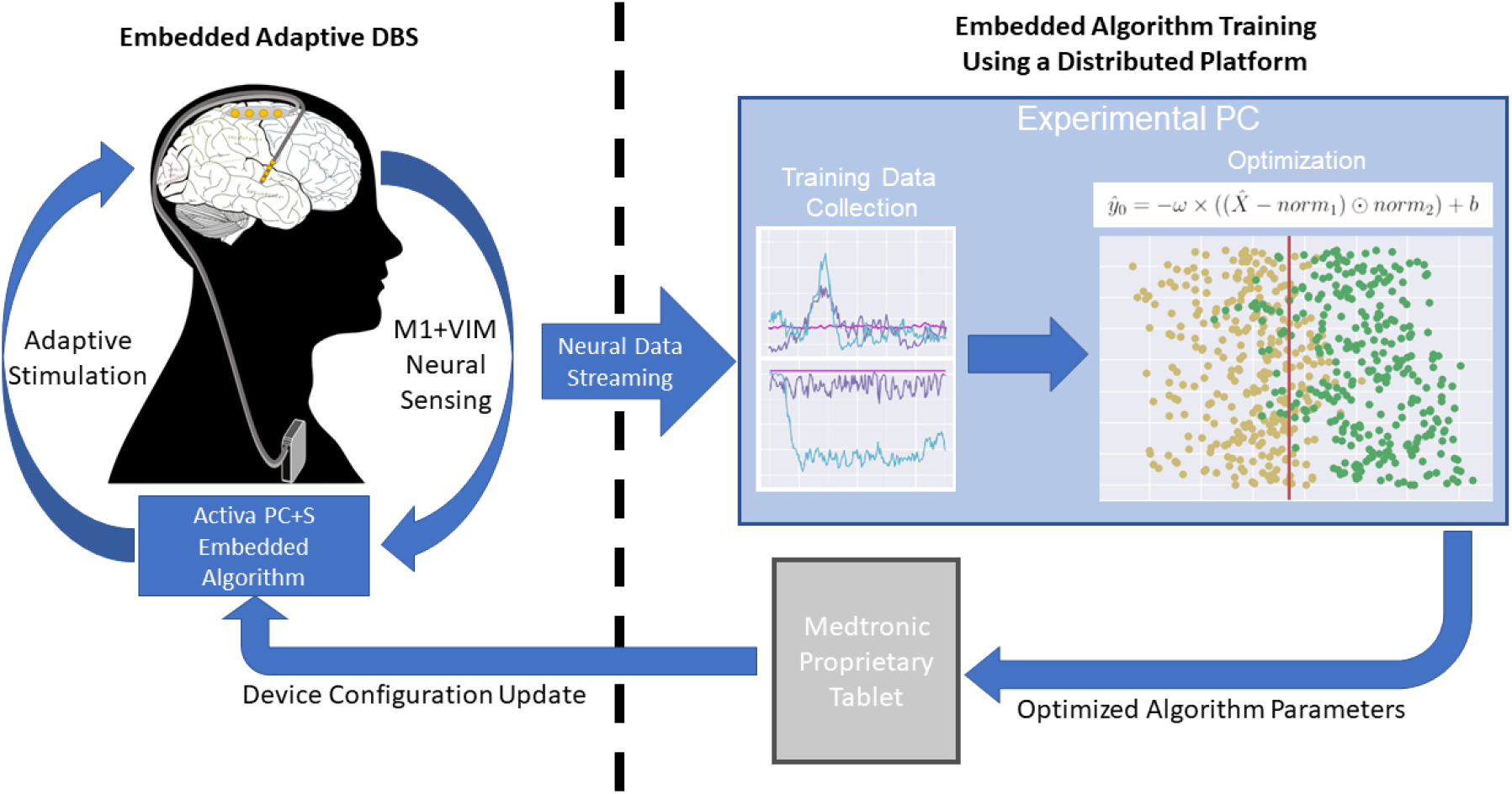
A diagram of our distributed training procedure using the Medtronic Activa PC+S. During the training procedure outlined in section 2.3, neural data is streamed wirelessly to an experimental PC on which initial data review is conducted. When satisfactory data has been collected, optimized algorithm parameters are trained and transported via USB to a proprietary tablet device. A configuration update to the Activa PC+S itself supplies it with the trained, personalized classifier. The embedded aDBS system then uses its sensing capacity with this classifier to modulate stimulation.

### 2.3. Supervised training data collection

Since only neural data may be used in our classifier and the presence of stimulation is known to alter neural dynamics, data must be collected in each state with stimulation both active and disabled. In order to take these considerations into account, training data must be collected with patients in each state of the form [*Stimulation State, Patient State*], amounting to A) [*Off, Rest*], B) [*Off, Action*], C) [*On, Rest*], and D) [*On, Action*].

To obtain data during each of the four possible patient states described above, 30 seconds of data were collected with the patient at rest with hands in lap with stimulation active and with stimulation disabled, and 30 seconds while the patient was continuously conducting the finger-to-nose task of the Fahn-Tolosa-Marin (FTM) tremor rating scale with stimulation active and with stimulation disabled [25]. These two minutes’ total data were used to train an intrinsically personalized classifier. Immediately following data collection, a visualization of the time series of the bandpower estimate data akin to that seen in fig. 2, along with cross-validated accuracy of a classifier trained on this data, was available for immediate review. This would inform the researchers’ decision to repeat individual tests if necessary. The structure of the training data collection process was arranged specifically to allow for individual states to be recorded independently, as opposed to necessitating a complete repetition of the full training procedure.

**Figure 2.**
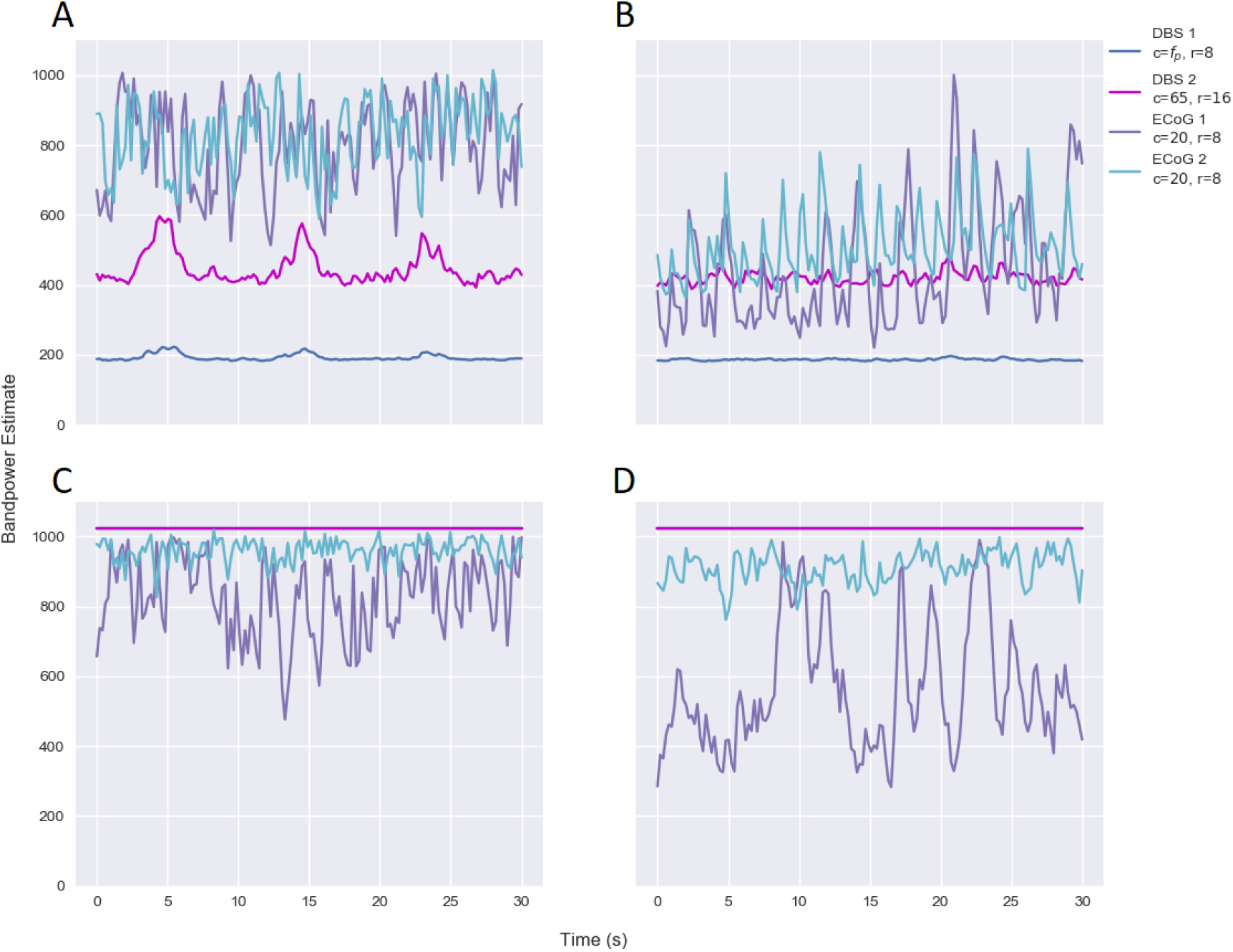
Time series training bandpower estimates of neural data used for Patient 1 for recordings, of form *[Stimulation State, Patient State*], of states A) [*Off, Rest*], B) [*Off, Action*], C) [On, Rest], and D) [On, *Action*]. Channel legend in upper right. Each set is 30 seconds in length, with bandpower estimate being an integer value between 0 and 1023. Differences between the *Rest* and *Action* states are readily apparent for ECoG data within stimulation states, with clear differences between the behavior across stimulation states. In the DBS channels, note the differences between signal behavior during *Rest* and *Action* with stimulation *Off*, and that both are saturated when stimulation is *On*. This implies that the DBS channels provide both an effective indication of whether stimulation is active and useful information when it is not.

### 2.4. Band selection and algorithm design

The primary task of the algorithm is to differentiate between when stimulation must be set to “HIGH”, and when it may be left “LOW”. The objective is therefore to determine when a patient requires stimulation to treat their symptoms, which in ET patients may be said to be the difference between when the patient is at *Rest* and generally without tremor, and when they are in *Action* and thus experiencing tremor.

Additionally, as the classifier has no explicit way to “know” whether stimulation is active or disabled, some method of indirect inference for device state must be provided. To allow the algorithm to determine whether stimulation was active or disabled, *x*_*DBS*,1_ bandpower estimate was set to the patient-specific, clinically determined stimulation frequency, (*c* = *f_patient_*, *r* = 8). *x*_*DBS*_,_2_ bandpower estimate was set to measure thalamic *γ*-band (*c* = 65, *r* = 16), previously demonstrated to correlate with movement [26]. Although suppression of thalamocortical coupling between the *γ*-band of the cortex and lower frequency bands of the VIM have been demonstrated to strongly correlate with movement [27], the noise floor in the Activa PC+S ECoG strip precludes effective measurement of this range. Instead, both *x*_*ECoG*,1_ and *x*_*ECoG*,2_ were set to record *β*-bandpower (*c* = 20, *r* = 8) from alternating pairs of the 4 linearly arranged ECoG electrodes available, desynchronization of which is known to correlate with movement onset, thus indirectly indicating onset of tremor in ET [20, 28, 29].

With 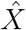 thus defined, an unbiased linear classifier was trained to differentiate between the 60 seconds of data collected during the *Rest* recordings and the 60 seconds collected during the *Action* recordings. A linear projection 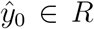 was defined with *ω* ∊ *R*^1×4^, *b* ∊ *R*, *norm*_1_ ∊ *R*^4×1^, and *norm*_2_ ∊ *R*^4×1^ of the form

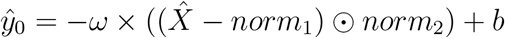

so as to maximize the variance between of 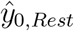 and 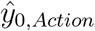. A sample above 0 was classified as *Action* and below 0 as *Rest*.

Finally, under the clinically informed theory that it is preferable to have stimulation unnecessarily active than to risk it being absent when needed, the classifier was biased in favor of keeping stimulation *On* by 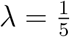 the standard deviation of the projection from training data. This was accomplished by adding *λ* to that projection, thus creating the final projection 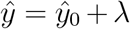. This fraction was determined through analysis of previously recorded data for our aDBS systems and found to consistently reduce false negatives without excessively increasing the overall error rate.

### 2.5. Tremor severity characterization and analysis

Gyroscope data from each task was extracted from IMU data streamed during testing, a 4 – 12Hz bandpass filter applied to extract only tremor-related data [2], and Welch’s method was applied to *x*, *y*, and *z* components individually. The area under the curve of each component was approximated with the trapezoidal method and the sum of these values used as our ground truth for semi-instantaneous IMU-based tremor severity assessment, denoted *χ*. Average level of tremor *S*, normalized for duration of test *τ* and thus defined as

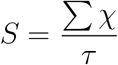

was derived for each test state. Tremor suppression *J* was defined as the fraction of tremor severity reduction as compared to tremor with stimulation *Off*, such that tremor suppression for a given control system may be defined

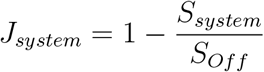

### 2.6. Quantifying reduction in energy use

Energy use per unit time was calculated using the total electrical energy delivered (TEED) methodology proposed by Koss and colleagues [30] adapted to control for test duration *τ*, such that

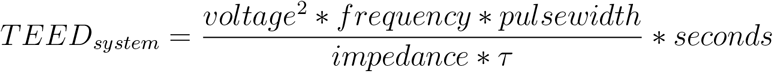

Energy use with aDBS will therefore be defined as the percentage of TEED saved in aDBS versus cDBS, such that relative energy saved 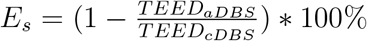

### 2.7. Methods for evaluation of therapeutic accuracy

For the quantified portion of classifier evaluation, patients were asked to begin at rest with hands in their lap. At a semi-randomized time-stamped prompt, the patient was asked to conduct the finger-to-nose task of the FTM tremor rating scale [25] continuously until the next prompt, at which point they were asked to return to rest. IMU data was streamed continuously throughout the experiment, while stimulation amplitude was recorded on the Activa PC+S device during experiments and downloaded for analysis following the experiment.

A false positive *ϵ*_+_ was said to occur when stimulation amplitude rose above 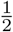 of a subject’s clinically prescribed settings during a rest period, while a false negative *ϵ*_−_ was defined as stimulation amplitude below this level during a period of movement. Total error rate *ε* was defined as the total number of errors divided by the duration of test *τ*, such that

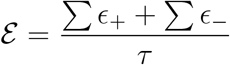

This protocol was conducted with stimulation disabled, cDBS control system active, and aDBS control system active.

Following this controlled experimental protocol patients were asked to stand and move freely for some minutes in order to determine qualitatively whether they could detect a difference in the quality of their treatment or in the manifestation of side effects.

## 3. Results

### 3.1. Performance of distributed procedure

As described in section 2.3, the data collected was made immediately available for review following recording of all 4 states. In each patient, review of the data visualization and cross-validated classifier performance led to the determination that one state, different in each patient, was excessively noisy. The individual test pertaining to that state was repeated, and the classifier trained from the new set of states uploaded to the device.

### 3.2. Performance of biased classifier on training data

For the training data, a false positive is said to have occurred if a sample labelled *Rest* is above the threshold, while a false negative is said to have occurred if a sample labelled *Action* is below the threshold. Total error rate is the average of these rates, as equal training time was spent in the *Rest* and *Action* states. Biasing the threshold resulted in a 12.7% increase in average error rate; however, this constituted a 28.2% decrease in average false negative rate. Training data distribution and classification in each patient are presented in fig. 3, and the full results of biasing are detailed in table 1.

**Figure 3.**
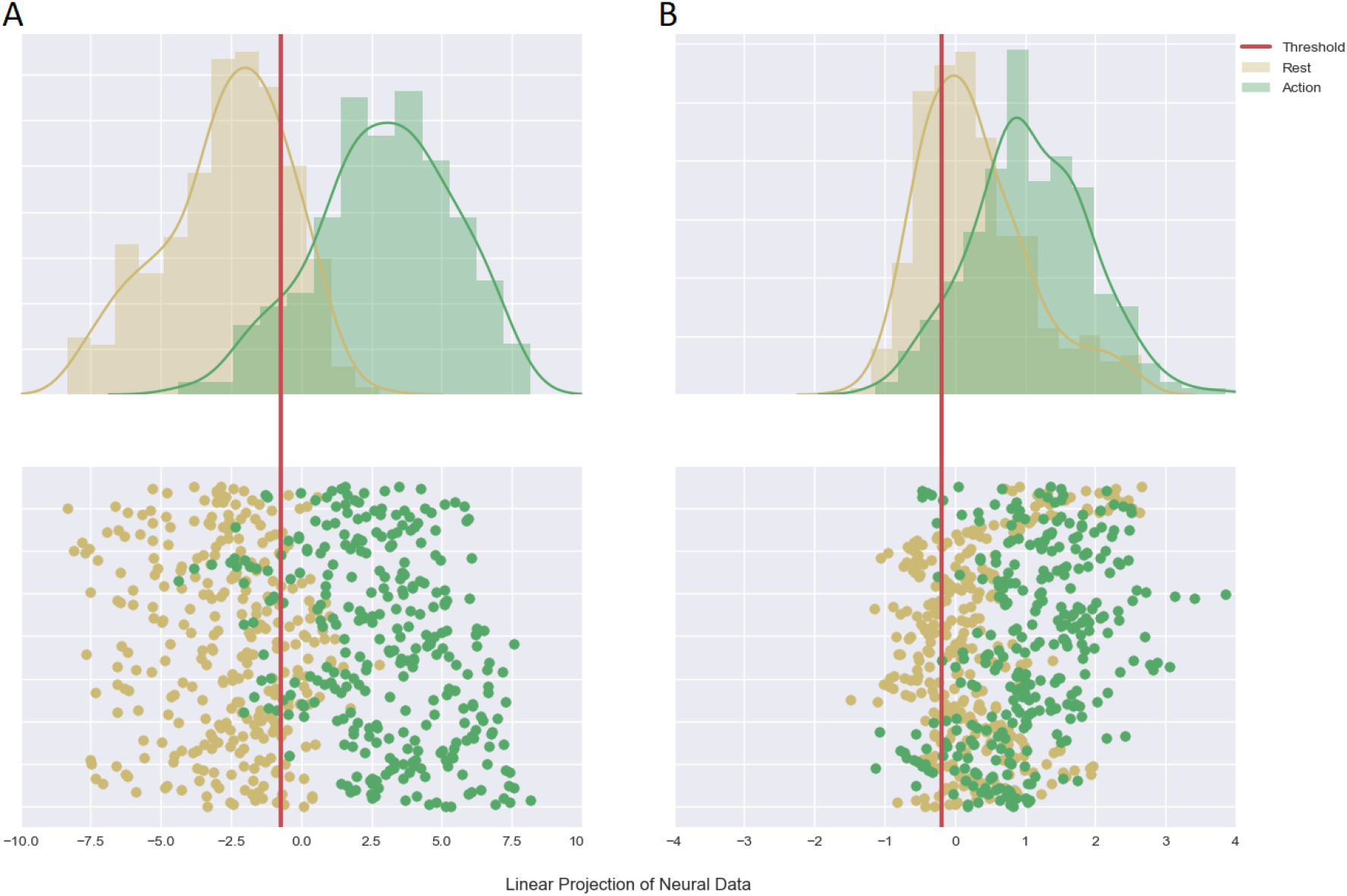
Kernel density estimation with histogram (top) and scatter plot of 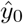 (bottom) in training data for A) Patient 1 and B) Patient 2, with *Rest* data for both stimulation settings combined in yellow and *Action* data for both stimulation settings combined in green. The unbiased classifier control system would return *On* for any point over 0 and *Off* for any point beneath 0. Our patient-specific biased threshold, seen here as a red line down each figure, demonstrates that appreciably more of the *Action* state is classified as requiring stimulation than the unbiased system would recognize.

**Table 1.** Effects of biasing on training data. Though overall error rate is increased by a marginal amount, clinically relevant false negative rate is substantially reduced in both patients, indicating that biasing was an effective method for increasing treatment reliability by clinical considerations.

| Measure | Unbiased | Biased |
| --- | --- | --- |
| P1 training error rate | 0.122 | 0.143 |
| P2 training error rate | 0.342 | 0.380 |
| P1 training false neg. rate | 0.130 | 0.090 |
| P2 training false neg. rate | 0.097 | 0.073 |

### 3.3. Therapeutic performance during controlled testing

For testing data, it was determined that therapeutic classifier average total error rate was *ε* = 0.468. However, over 92% of these errors were comprised of clinically permissible false positives; average false negative rate was *ε*_−_ = 0.036, indicating that stimulation was almost always being supplied at therapeutic levels during our experimental procedure. Use of the aDBS control system resulted in a 30.8% average drop in energy use by the neurostimulator. Complete results may be found in table 2, with stimulation and tremor severity during experiments displayed in fig. 4.

**Figure 4.**
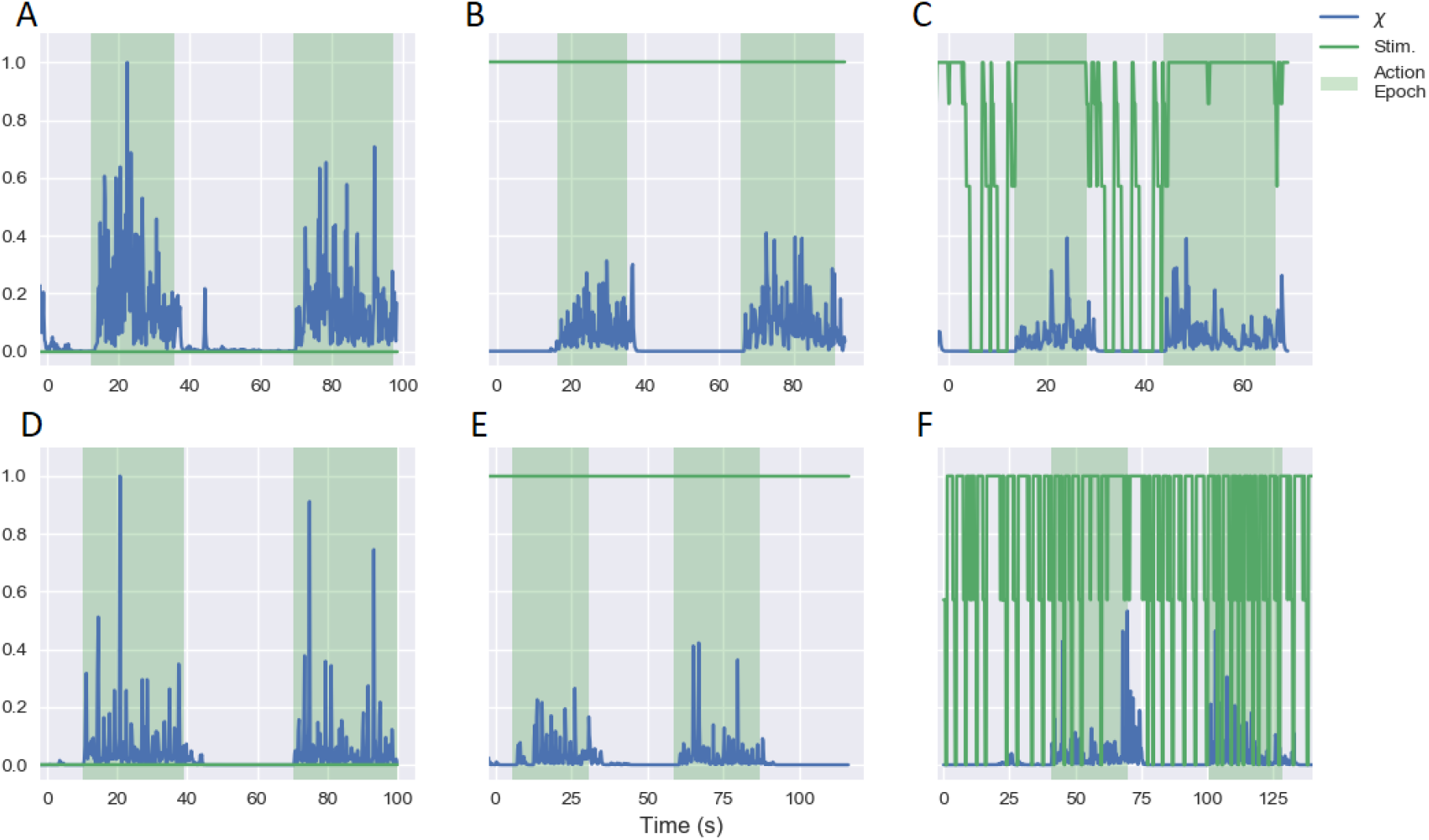
Stimulation amplitude (green line), normalized by clinically determined maximum amplitude, and tremor severity *χ* (blue line), normalized to the maximum value in any state, with stimulation disabled, enabled and with aDBS active (respectively, A, B, and C in Patient 1 and D, E, and F in Patient 2). Green background indicates patient was asked to perform finger-to-nose task, while blank background indicates patient instructed to rest with hands in lap.

**Table 2.** Efficacy of aDBS. The extremely low false negative rates, paired with a significant reduction in overall stimulation, appear to have led to a substantial increase in tremor suppression in aDBS over cDBS. The mechanism for this is not well understood at present.

| Patient | 1 | 2 |
| --- | --- | --- |
| Percent energy saved | 34.0% | 27.5% |
| Overall error rate | 0.361 | 0.575 |
| False negative rate | 0.030 | 0.042 |
| cDBS tremor suppression | 0.596 | 0.125 |
| aDBS tremor suppression | 0.740 | 0.221 |

During their free movement period, subjects clearly and accurately differentiated between when stimulation was disabled and when some stimulation control system was active. However, they reported no differences in treatment efficacy between aDBS and cDBS. One patient reported experiencing a brief paresthesia in their right hand on two occasions with aDBS active. Neither reported any side effects with cDBS active.

### 3.4. Tremor suppression in cDBS vs aDBS

Average tremor suppression across both patients with cDBS *J_cDBS_* = 0.361. Average tremor suppression with aDBS *J_aDBS_* = 0.481 for a 33.2% improvement with aDBS over cDBS. This is in line with previous findings that aDBS is more effective in tremor suppression than cDBS [14, 20].

## 4. Discussion

### 4.1. Data collection and classifier training

The versatility of our distributed training system ensured that the entirety of the procedure, including explaining procedure and tests to subject, recording training data, reviewing data and repeating tests if needed, training classifier on selected data sets, and uploading classifier to patient device, was completed in under 20 minutes in each patient. This training time included repeat data collection for one state in each patient when initial data review revealed insufficient classification accuracy. This indicates that the brevity of the training process was due largely to the rapidity with which data could be analyzed and the ease of collecting more at-will with the distributed system.

### 4.2. Effectiveness of biased classification

Biasing the classifier towards the *On* state reduced overall accuracy; however, this was exactly the intent. As may be seen in the analysis of training data, biasing resulted in marginal increases in overall error rate while cutting false negative rates by 28.7% for an overall sensitivity of 91.8%. In our analysis of therapeutic accuracy during testing, we found that 96.4% of the time stimulation was required, it was provided. This increase in sensitivity may have to do with alteration of neural dynamics during stimulation ramping periods. Although the effect appears to be an increase in the likelihood of an *On* control signal, giving little clinical reason to examine it more closely, it is nonetheless an interesting occurrence that may be considered further in future works.

### 4.3. Tremor reduction during quantified testing

In keeping with previous findings for ET [14, 20], our quantified analyses of symptom severity indicated that aDBS was substantially more effective in tremor suppression than cDBS on average, albeit with less substantial tremor reduction in P2 than in P1. This divergence may be partly due to P1 having considerably more severe tremor without treatment, reducing the amount that treatment can effectively reduce symptoms in P2. This may be similar to the “flooring” effects found in [18]. One follow up question with larger patient pools will be whether aDBS is as much more effective in all patients, whether it improves tremor suppression in some individual patients more than others, or if it is simply more effective at treating certain conditions, such as ET vs PD.

### 4.4. aDBS in freely moving patients

That aDBS treatment was at least as effective as cDBS was supported by our patients’ reports in their periods of free movement, during which they noticed no substantial differences in treatment efficacy. Paired with our quantitative results implying aDBS may be more effective than cDBS, this suggests that, while the differences in therapeutic efficacy may be below the threshold of perception for most patients already receiving cDBS, an aDBS system may be able to operate with a lower maximum amplitude than that used in cDBS systems, thereby reducing overall stimulation to an even greater extent. This will further increase the already substantial energy savings promised by aDBS systems.

The transient paresthesia experienced by Patient 1 is most likely an indicator that the ramping rate for stimulation was set to too high a level [15]. This issue is straightforward to resolve during the patient programming phase. A “maximum tolerable rate” test will be implemented in future training procedures in order to ensure patient comfort is maintained during aDBS.

## 5. Conclusion

Here we have demonstrated one of the first examples of a fully implanted aDBS system in freely moving ET patients, and the first with a distributed training procedure and biased classification algorithm. In addition to demonstrating the efficacy of this system, our evidence supports earlier work indicating that aDBS is quantifiably more effective than cDBS in experimental settings and results in no substantial difference in treatment efficacy from the patient’s perspective. The combination of increased overall efficacy, a novel, translation-ready training procedure, and a clinically-oriented classification algorithm indicate that fully implanted aDBS systems are approaching a level of clinical efficacy needed for population-wide deployment. This validation in clinical settings will serve as the base for chronic, at-home aDBS studies in the near future, the evaluation of which is one of the last barriers to adoption of aDBS as a new standard of care for an era of personalized medicine.

## Data Availability

Data available only at the sole discretion of the senior authors.

## Acknowledgments

The authors wish to acknowledge Timothy Brown for innumerable ethical insights that have informed the structure of these experiments and Medtronic for technical and material support.

